# Performance of GPT-4V(ision) in Ophthalmology: Use of Images in Clinical Questions

**DOI:** 10.1101/2024.01.26.24301802

**Authors:** Kosei Tomita, Takashi Nishida, Yoshiyuki Kitaguchi, Masahiro Miyake, Koji Kitazawa

## Abstract

**Background/aims:** To compare the diagnostic accuracy of Generative Pre-trained Transformer with Vision (GPT)-4 and GPT-4 with Vision (GPT-4V) for clinical questions in ophthalmology.

**Methods:** The questions were collected from the “Diagnosis This” section on the American Academy of Ophthalmology website. We tested 580 questions and presented GPT-4V with the same questions under two conditions: 1) multimodal model, incorporating both the question text and associated images, and 2) text-only model. We then compared the difference in accuracy between the two conditions using the chi-square test. The percentage of general correct answers was also collected from the website.

**Results:** The GPT-4V model demonstrated higher accuracy with images (71.7%) than without images (66.7%, p<0.001). Both GPT-4 models showed higher accuracy than the general correct answers on the website [64.6 (95%CI, 62.9 to 66.3)].

**Conclusions:** The addition of information from images enhances the performance of GPT-4V in diagnosing clinical questions in ophthalmology. This suggests that integrating multimodal data could be crucial in developing more effective and reliable diagnostic tools in medical fields.

**SYNOPSIS:** The study compared the diagnostic accuracy of GPT-4 and GPT-4 with Vision for clinical questions in ophthalmology, finding that the performance improved when it analyzed both text and images.

**WHAT IS ALREADY KNOWN ON THIS TOPIC:** Text-based large language models (LLMs) have demonstrated significant potential in enhancing medical interpretation and diagnosis. Generative Pretrained Transformer 4 with Vision (GPT-4V) can address image-related questions, but the use of GPT-4V in ophthalmology has not yet been validated.

**WHAT THIS STUDY ADDS:** Our study reports the answer accuracy on ‘Diagnose This,’ provided by the American Academy of Ophthalmology, using GPT-4V. The integration of image data with GPT-4V enhances diagnostic accuracy in addressing ophthalmic clinical questions.

**HOW THIS STUDY MIGHT AFFECT RESEARCH, PRACTICE OR POLICY:** Our study indicates that combining image data with GPT-4 can enhance diagnostic accuracy in ophthalmic clinical questions. The development of LLMs trained on medical-specific datasets could further increase accuracy, advancing towards practical clinical applications.

## Introduction

The field of natural language processing has developed rapidly with the advent of large language models (LLMs).[1] Text-based LLMs have shown promise in enhancing medical interpretation and diagnosis. Specifically, in ophthalmology, the efficacy of these models has been explored in various exams, including the Basic and Clinical Science Course (BCSC) Self-Assessment Program, OphthoQuestions question banks, and FRCOphth examinations.[2–4] Furthermore, it is important to note that LLMs are evolving and the performance of the model may improve dramatically in a short period of time.

OpenAI recently introduced Generative Pre-trained Transformer (GPT)-4V(ision), a robust multimodal model excelling in image interpretation. GPT-4V is a versatile LLM that can process both images and text, facilitating tasks such as visual question answering.[5] GPT-4V relies on image understanding for visual question answering (VQA), a field integrating computer vision and natural language processing.[6] This integration allows GPT-4V to analyze and understand visual data, enhancing its capability to interpret and respond to text. Its training data includes a vast array of texts and images that were available up to April 2023. However, it does not directly compare a specific image with another previously published image. GPT-4V has the capability to respond to image-related queries. The introduction of multimodal LLM that can process diverse inputs, including images, exhibits a significant advancement.[7] Incorporating medical images into these multimodal LLMs could potentially improve their efficacy in addressing clinical questions.

This study aims to evaluate and compare the diagnostic accuracy of text-based and multimodal LLMs in ophthalmology, thereby deepening our understanding of advanced AI models in healthcare, especially in image-centric disciplines like ophthalmology. It has the potential to improve AI-assisted diagnostics in eye care, benefiting both practitioners and patients.

## Methods

In our study, we employed the latest version of the language model available at the time of study (GPT-4 turbo, OpenAI; https://chat.openai.com/), which is a multimodal model capable of processing both text and images. This version of ChatGPT was trained with information available up to April 2023. We performed the following prompt: “I am conducting an experiment on ophthalmic clinical case discussions to compare your diagnostic conclusions with those of clinicians. Each case is derived from the American Academy of Ophthalmology website. You are not trying to treat the patients. At the end of the case, you will be presented with four choices. Please select the most likely answer. If no background information or images are provided, please solve the question with only the available information. There’s no need to elaborate on your reasoning.” We tested the same questions under two conditions: 1) multimodal model, incorporating both the question text and associated images, and 2) text-only model. Text and images were manually entered via the ChatGPT web interface. To prevent the possibility of past responses affecting the output of ChatGPT, a new chat session was created for each question and each condition (i.e., with and without images). These procedures were conducted between December 18 and December 26, 2023.

The questions were collected from the “Diagnosis This” section on American Academy of Ophthalmology (AAO) website (https://www.aao.org/education/education-browse?filter=diagnose-this). Since November 2009, the “Diagnosis This” program has presented one case almost every week, accumulating a total of 677 cases. Each case consists of both image and text, and one answer is chosen from four options. The order of these options is randomized on the website, and subsequently, the order of the four options entered into the prompt is also randomized. After responding, the correct answer and explanation for the case, and the rates for general correct answers are displayed on the website. The purpose of this study was to compare the performance of multimodal models and text-only models; therefore, the uses of non-specific images (n=57) were excluded.

Furthermore, lack of answers from the text model (n=27), presence of duplicated questions (n=8), absence of four options for the question (n=4), and unavailability of the question on the website (n=2) were also excluded. Consequently, 580 questions were included in the dataset (**Figure 1**). The subspecialty in question was also recorded.

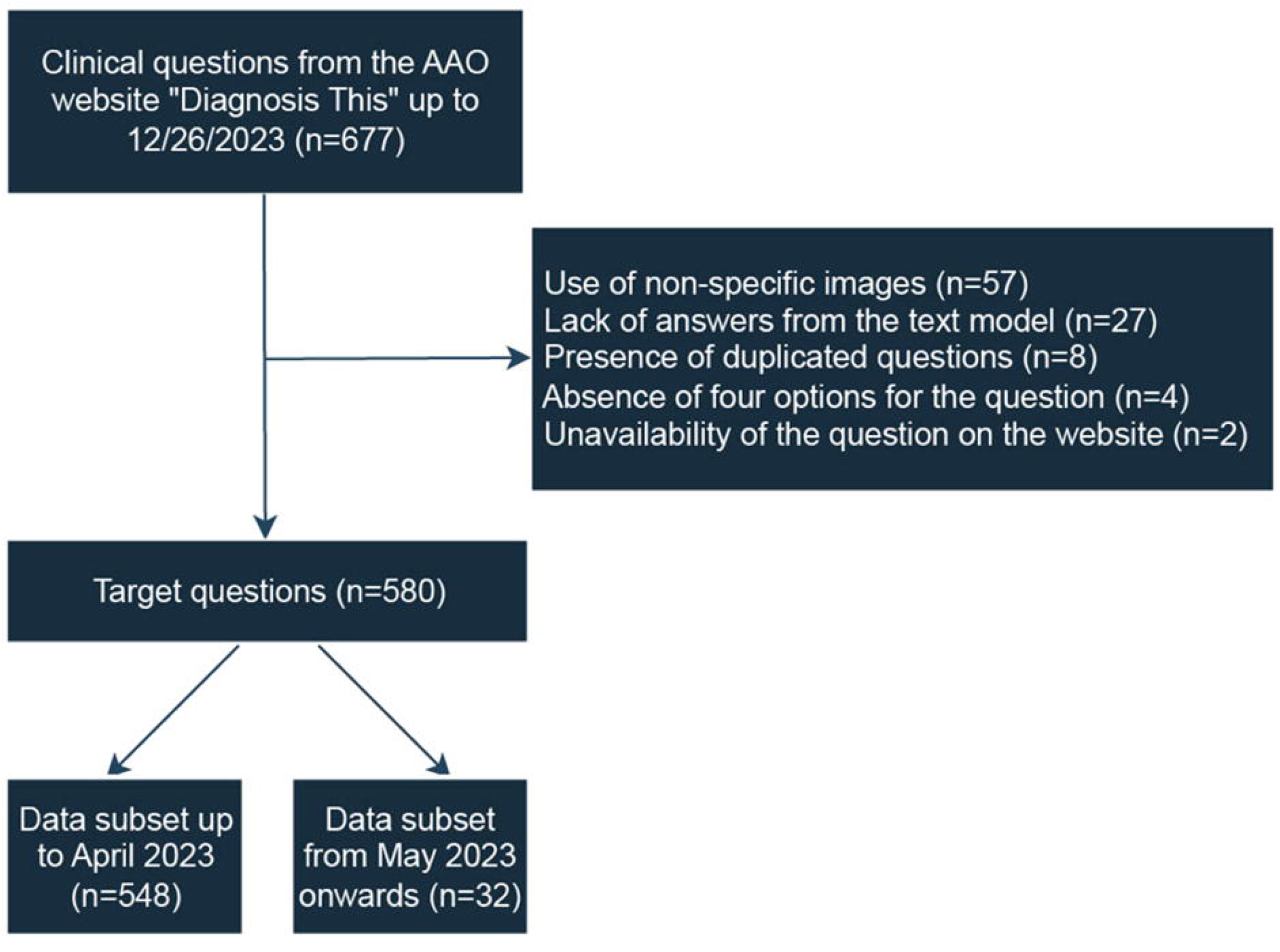

It has not been disclosed whether the AAO website is included in the training data for ChatGPT. Therefore, to avoid the influence of training on the responses, subsets of data up to April 2023 and from May 2023 onwards were created and compared. Furthermore, for the subset after May 2023, prompts were repeated five times for both the multimodal and text models, with the order of the multiple-choice options randomized, and an intraclass correlation coefficient (ICC) was calculated to assess the reproducibility of the correct answers.

Chi-square tests were conducted to compare the correct answer rate between multimodal and text-only models. Statistical analyses were performed using Stata version 16.0 (StataCorp, College Station, TX) and python 3.11.1 (Python Software Cooperation, Wilmington, DE, USA). There were no corrections of P values made for multiple comparisons. All P values were two-sided. This research did not involve human subjects, and therefore, did not require Institutional Review Board approval.

## Results

The accuracy was 71.7% for the multimodal model and 66.7% for the text-only model (p<0.001; **Table 1**). The results for the subset divided by the end of April 2023 showed a similar trend (**Supplemental Table 1**). ICC was 0.91 (95%CI, 0.87 to 0.94) for the data subset from May 2023 onwards.

**Table 1.** Accuracy of multimodal model (with images) and text only model (without images)

| Overall ($P<0.001$) | | Multimodal model (with images) | | |
| --- | --- | --- | --- | --- |
|  |  | Correct | Incorrect | Total |
| Text only model (without images) | Correct | 355 (61.2%) | 32 (5.5%) | 387 (66.7%) |
|  | Incorrect | 61 (10.5%) | 132 (22.8%) | 193 (33.3%) |
|  | Total | 416 (71.7%) | 164 (28.3%) | 580 (100.0%) |

**Table 2** shows the accuracy of the multimodal model, text model, and rate of general correct answers on the website. The multimodal model [71.7 (95%CI, 68.0 to 75.4)] showed higher diagnostic accuracy compared to the text-only model [66.7 (95%CI, 62.9 to 70.6)] and the general correct answers on the website [64.6 (95%CI, 62.9 to 66.3)].

**Table 2.** Accuracy of multimodal models, text models, and general correct answers on the website.

| Correct | Overall | Subset up to 4/2023 | Subset from 5/2023 |
| --- | --- | --- | --- |
| Multimodal models (with images) | 71.7 (68.0 to 75.4) | 71.9 (55.4 to 88.3) | 71.9 (55.4 to 88.3) |
| Text only model (without images) | 66.7 (62.9 to 70.6) | 68.8 (51.8 to 85.7) | 68.8 (51.8 to 85.7) |
| General correct answers on the website | 64.6 (62.9 to 66.3) | 64.7 (56.8 to 72.6) | 64.7 (56.8 to 72.6) |

Values are shown in mean (95% confidence interval).

**Figure 2** shows the percentage of correct answers for each subspecialty. The accuracy rate was generally around 60-80% across various subspecialties. However, in the categories of retina and vitreous (n=84) with a correct rate of 76.2%, oculofacial plastic and orbital surgery (n=75) with 87.8%, and glaucoma (n=48) with 79.2%, the multimodal model demonstrated a higher accuracy rate compared to the text-only model and the general correct answers on the website.

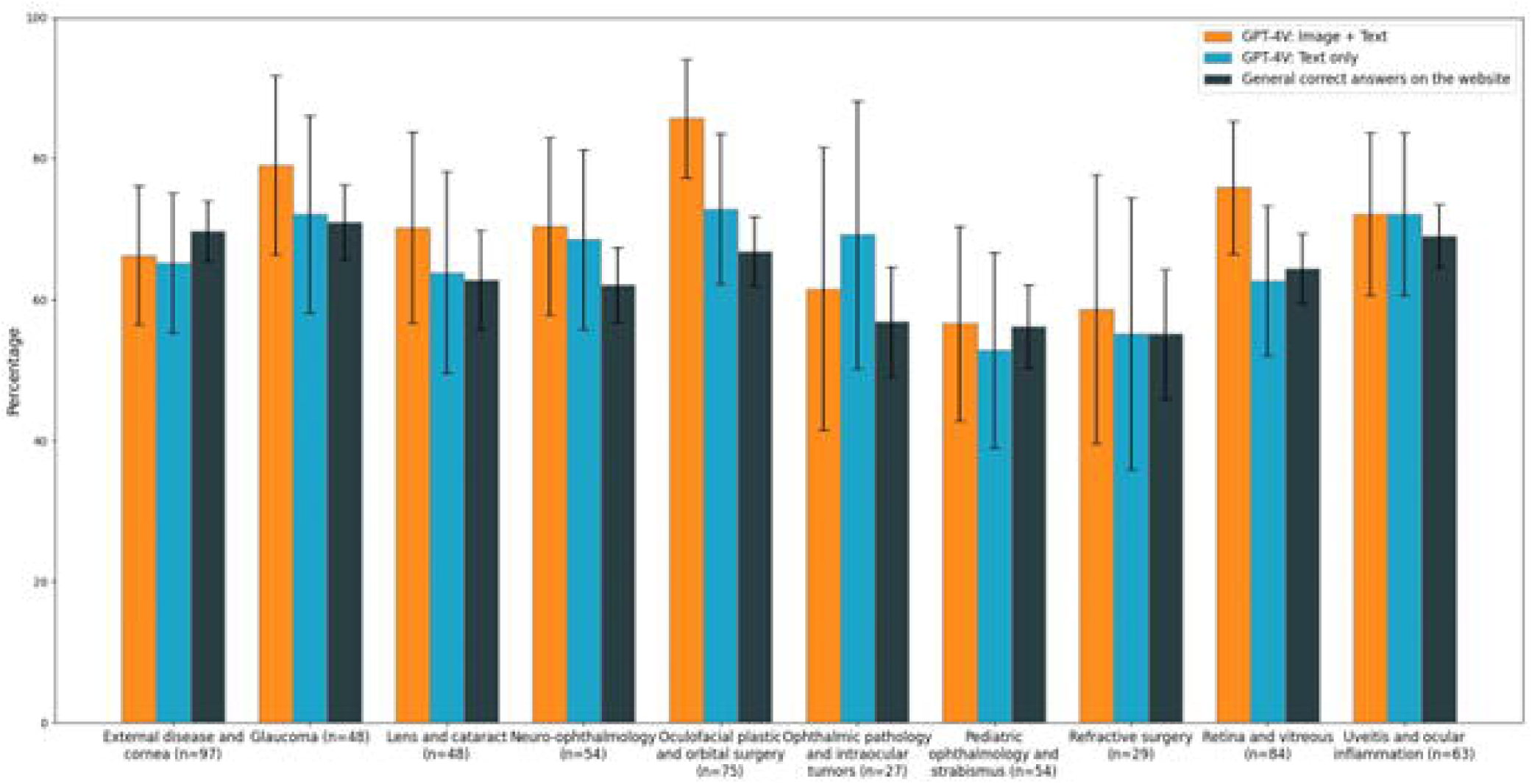

## Discussion

In the current study, we compared the diagnostic accuracy of GPT-4-based text model and multimodal model for ophthalmic clinical questions. By incorporating information from images in addition to text, the performance of GPT-4V in diagnosing clinical questions improved. The improvement was notable in oculofacial plastic and orbital surgery, retina and vitreous, and glaucoma. Despite significant benefits, the overall accuracy rate was not consistently high, indicating that there are still hurdles to its clinical use. Nevertheless, GPT-4V outperformed general respondents in clinical question accuracy, highlighting the potential for future developments in LLMs.

Previous studies have explored the accuracy of LLMs in answering standardized text-only ophthalmology questions. One such study randomly selected 260 text-only questions from BCSC and OphthoQuestions, adjusted for the level of cognition and question difficulty to investigate the accuracy of GPT-4. The finding indicated a combined accuracy rate of 72.9%.[4] In the current study, the accuracy rate of GPT-4V on ‘Diagnose This,’ for the text model was 66.7%. In our current study, the accuracy rate for the text model was slightly lower compared to the previous studies. This discrepancy may result from our study not adjusting for difficulty levels, leading to considerable variation in the difficulty of the problem statements. Even under these conditions, the increase in the accuracy rate for multimodal model to 71.7% may suggest its capability to correctly answer more challenging questions. For instance, in a case presenting central visual loss, a full-thickness macular hole was diagnosed from optical coherence tomography (OCT) and fundus photograph, leading to a correct treatment plan. This question was misanswered with text-only model. In this study, GPT-4V demonstrated proficiency in processing various medical images and identifying specific features, yet it is important to note that the model occasionally failed to recognize overt findings. There is a case, like the misidentification of acute corneal hydrops in keratoconus as Acanthamoeba keratitis, indicating areas for further improvement (Supplemental Result 1).

Our study found that the overall accuracy rate of the multimodal model was higher than that of text-only model, with varying degrees of improvement across different subspecialties. In specific subspecialties, such as oculofacial plastic and orbital surgery, retina and vitreous, and glaucoma, accuracy rates notably increased, likely due to image availability. In ophthalmology, imaging is crucial for diagnosis and management in clinical practice. For example, glaucoma and retinal diseases are particularly reliant on OCT. Similarly, oculofacial plastic and orbital surgery questions often involved computerized tomography (CT) and magnetic resonance imaging scans. GPT-4V demonstrated proficiency in interpreting and responding to clinical imaging, suggesting that image availability may enhance response accuracy. In contrast, for external disease and cornea, and pediatric ophthalmology and strabismus, the use of imaging information did not improve accuracy. Recognizing complex structures such as the cornea, conjunctiva, and eyelids in images can result in inaccuracies. Strabismus diagnosis with images remains difficult, due to the complexity of eye muscle movements and surgical decision-making. There is a possibility that information related to medical imaging, such as OCT and CT scans, which significantly contribute to our clinical practice, is widely available and publicly accessible on the web. This suggests that sub-specialties utilizing these imaging resources might particularly benefit from the use of image-assisted LLMs. On the other hand, for subspecialties that did not readily benefit from the presence of images in this study, it may be possible to enhance overall answer accuracy in the future by providing specialized image training sets individually to these subspecialties.

LLMs are increasingly valuable as innovative tools for interpreting the visual world. They provide descriptions of photographs from smartphones for individuals who are blind or have low vision, detailing the surrounding environment, object locations, and character recognition. Ongoing efforts aim to employ these technologies in patient care, aiming to alleviate the workload of healthcare providers in clinical settings. Reported inaccuracies in responses highlight an area of concern.[8] OpenAI explicitly states that the current version of GPT-4V is unsuitable for any medical function, including providing professional medical advice, diagnoses, treatments, or judgments, due to its suboptimal performance in the medical field.[5] There have been cases where its use in interpreting medical images led to severe errors, such as incorrect identification of lesion laterality.[5,9] These issues highlight the need for further enhancements in model accuracy, validation, regulatory compliance, and ethical considerations before it can be safely applied in clinical contexts. Based on our findings, GPT-4V using multimodal model showed an increased accuracy compared to text-only model. However, when considering its immediate applicability in clinical settings, the 71.9% accuracy rate of GPT-4V is a nuanced result, being neither too high nor too low. The potential for LLMs in medicine is promising, yet it’s important to acknowledge that resources like BCSC, OphthoQuestions, and Diagnose This are ultimately structured as question texts, presumably with hints provided within the text for answering. In real clinical scenarios, the challenge extends beyond interpreting physical signs to accurately gathering patient history for effective treatment. Effective communication between healthcare professionals and patients is essential for precise information collection.[4] Without such interaction, research using LLMs might remain theoretical and not practically applicable, potentially delaying their real-world clinical implementation to a distant future. A systematic review highlights the growing anticipation for patient communication-focused LLMs, particularly in extracting patient information.[10] Looking ahead, the development of medically specialized LLMs is anticipated, alongside advancements in patient communication capabilities.

Effective use of LLMs in information gathering depends on the user’s understanding and the quality of prompts provided, therefore, effective collaboration between users and LLMs is crucial. Future advancement is expected to produce more sophisticated LLMs, driven by expanded training in specific domains. However, generally available LLMs may not yet be adequately trained on medical images due to the limited availability and privacy restrictions associated with such data. While LLMs benefit from extensive web text data, accessing medical imagery is challenging. Some researchers are now focusing on creating specialized multimodal LLMs for medical applications, utilizing open-source technologies and public resources.[11–13] Despite the scarcity of medical images online, those from electronic medical records, paper-based documentation, and the scientific articles could serve as valuable training datasets for these more focused multimodal LLMs. The development of domain-specific models holds considerable promise in specialized areas like medicine.[2] LLM landscape has recently expanded with several models, not limited to ChatGPT, capable of processing images. Some of these models are noted for their superior performance. However, this study did not incorporate these recent other models. Given the rapid evolution of the field, it is foreseeable that more sophisticated models with enhanced capabilities will soon be developed. Integrating these models could facilitate more detailed and nuanced clinical assessments.

In conclusion, this study demonstrates that incorporating image data with GPT-4V improves diagnostic accuracy in ophthalmic clinical problems, indicating the potential of multimodal LLMs in medical applications. However, challenges such as AI hallucinations, errors, and limitations related to model design and policy, alongside the occasional failure in interpreting medical images, underscore the need for caution and further enhancements before these technologies can be safely implemented in clinical settings. Despite these challenges, the evolving landscape of LLMs, including the development of specialized multimodal models for medicine, holds promise for more sophisticated and nuanced clinical assessments in the future.

## Contributors

Involved in design and conduct of study: KT and TN, Data collection: KT and TN, Analysis and interpretation of data: TN, Writing: KT and TN, Critical revision: all authors, Approval of the manuscript: all authors, Guarantor of work: TN.

## Disclaimer

Not applicable

## Patient consent for publication

Not applicable.

## Ethics approval

No human subjects were included in this study. This study did not require institutional review board approval.

## Data availability statement

The datasets generated and/or analysed during the current study are available from the corresponding author on reasonable request.

## References

1 Introducing ChatGPT. [online]. 2023. https://openai.com/blog/chatgpt (accessed 17 January 2023).

2 Tan TF, Thirunavukarasu AJ, Campbell JP, et al. Generative Artificial Intelligence Through ChatGPT and Other Large Language Models in Ophthalmology. Ophthalmology Science. 2023;3:100394. doi:10.1016/j.xops.2023.100394

3 Antaki F, Touma S, Milad D, et al. Evaluating the Performance of ChatGPT in Ophthalmology: An Analysis of Its Successes and Shortcomings. Ophthalmology science. 2023;3. doi:10.1016/J.XOPS.2023.100324

4 Antaki F, Milad D, Chia MA, et al. Capabilities of GPT-4 in ophthalmology: an analysis of model entropy and progress towards human-level medical question answering. Br J Ophthalmol. 2023;bjo-2023-324438.

5 GPT-4V(ision) System Card. [online]. 2023. https://openai.com/research/gpt-4v-system-card (accessed 17 January 2023).

6 Jha S, Dey A, Kumar R, et al. A Novel Approach on Visual Question Answering by Parameter Prediction using Faster Region Based Convolutional Neural Network. International Journal of Interactive Multimedia and Artificial Intelligence. 2019;5:30–7. doi:10.9781/ijimai.2018.08.004

7 Zhang J, Huang J, Jin S, et al. Vision-Language Models for Vision Tasks: A Survey. arXiv:2305.12031 [Preprint]. 2023 10.48550/arXiv.2304.00685.

8 Sallam M. ChatGPT Utility in Healthcare Education, Research, and Practice: Systematic Review on the Promising Perspectives and Valid Concerns. Healthcare 2023, Vol 11, Page 887. 2023;11:887. doi:10.3390/healthcare11060887

9 Yang Z, Li L, Lin K, et al. The Dawn of LMMs: Preliminary Explorations with GPT-4V(ision). arXiv:2309.17421 [Preprint]. September 01, 2023 10.48550/arXiv.2309.17421.

10 Garg RK, Urs VL, Agarwal AA, et al. Exploring the role of ChatGPT in patient care (diagnosis and treatment) and medical research: A systematic review. Health Promot Perspect. 2023;13:183. doi:10.34172/hpp.2023.22

11 Toma A, Lawler PR, Ba J, et al. Clinical Camel: An Open Expert-Level Medical Language Model with Dialogue-Based Knowledge Encoding. arXiv:2305.12031 [Preprint]. 2023 10.48550/arXiv.2305.12031.

12 Singhal K, Azizi S, Tu T, et al. Large language models encode clinical knowledge. Nature 2023 620:7972. 2023;620:172–80.

13 Tu T, Azizi S, Driess D, et al. Towards Generalist Biomedical AI. arXiv:2305.12031 [Preprint]. July 02, 2023 10.48550/arXiv.2307.14334.

