## Supplemental for "Performance of GPT-4V(ision) in Ophthalmology: Use of Images in Clinical Questions"

**Supplemental Table 1.** Accuracy of multimodal model (with images) and text only model (without images) divided into subsets as of the end of April 2023

| **Subset up to 4/2023** (P<0.001) | | Multimodal model (with images) | | |
|---|---|---|---|---|
| | | Correct | Incorrect | Total |
| Text only model (without images) | Correct | 334 (61.0%) | 31 (5.7%) | 365 (66.6%) |
| | Incorrect | 59 (10.8%) | 124 (22.6%) | 183 (33.4%) |
| | Total | 393 (71.7%) | 155 (28.3%) | 548 (100.0%) |
| **Subset from 5/2023** (P<0.001) | | Multimodal model (with images) | | |
| | | Correct | Incorrect | Total |
| Text only model (without images) | Correct | 21 (65.6%) | 1 (3.1%) | 22 (68.8%) |
| | Incorrect | 2 (6.3%) | 8 (25.0%) | 10 (31.3%) |
| | Total | 23 (71.9%) | 9 (28.1%) | 32 (100.0%) |

**Supplemental Result**

The cases used in this study are sourced from questions available on the internet and are not intended to identify individual patients. Following the guidelines for submissions, some information, like the precise ages, has been modified in this supplemental document. For the original problem statements, please see the provided URL.

**Case 1:** A case with OCT and fundus photograph was accurately interpreted and answered correctly. The percentage for the general correct answer is 93.

https://www.aao.org/education/diagnose-this-player/diagnose-this-1-month-of-reduced-central-visual-fu

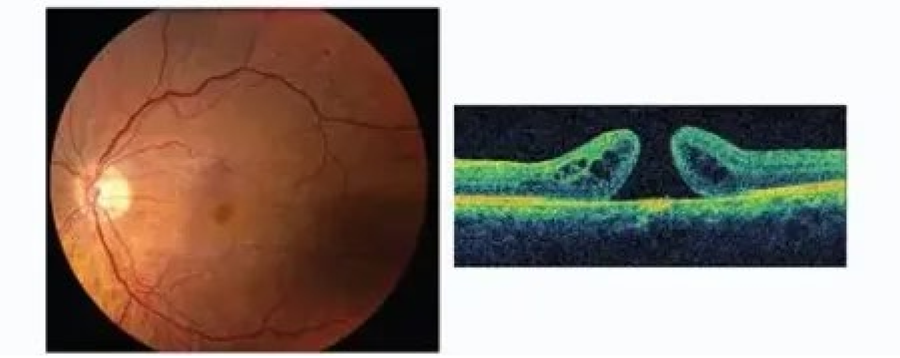

A woman in her 50s presents with 1 month of reduced central visual function in her left eye. Her ocular history is significant for presbyopia. Fundus and OCT are shown. What therapeutic intervention has the best prognosis for improvement of visual function in this patient?

1, Intravitreal ocriplasmin

2, Laser photocoagulation

3, Observation

4, Pars plana vitrectomy, posterior vitreous hyaloid/internal limiting membrane dissection, postop gas tamponade

Multimodal model (with images) answer: Based on the provided fundus photograph and Optical Coherence Tomography (OCT) image, and given the options for a woman in her 50s with reduced central visual function in her left eye for 1 month: The OCT image displays what appears to be a full-thickness macular hole. The best therapeutic intervention for a full-thickness macular hole is generally: Pars plana vitrectomy, posterior vitreous hyaloid/internal limiting membrane dissection, postop gas tamponade This procedure aims to relieve vitreomacular traction and to promote the closure of the hole, which can improve visual function.

**Case 2:** A case with an image information was accurately interpreted and answered correctly. The percentage for the general correct answer is 14.

https://www.aao.org/education/diagnose-this-player/diagnose-this-what-medication-is-most-likely-to-ca-2

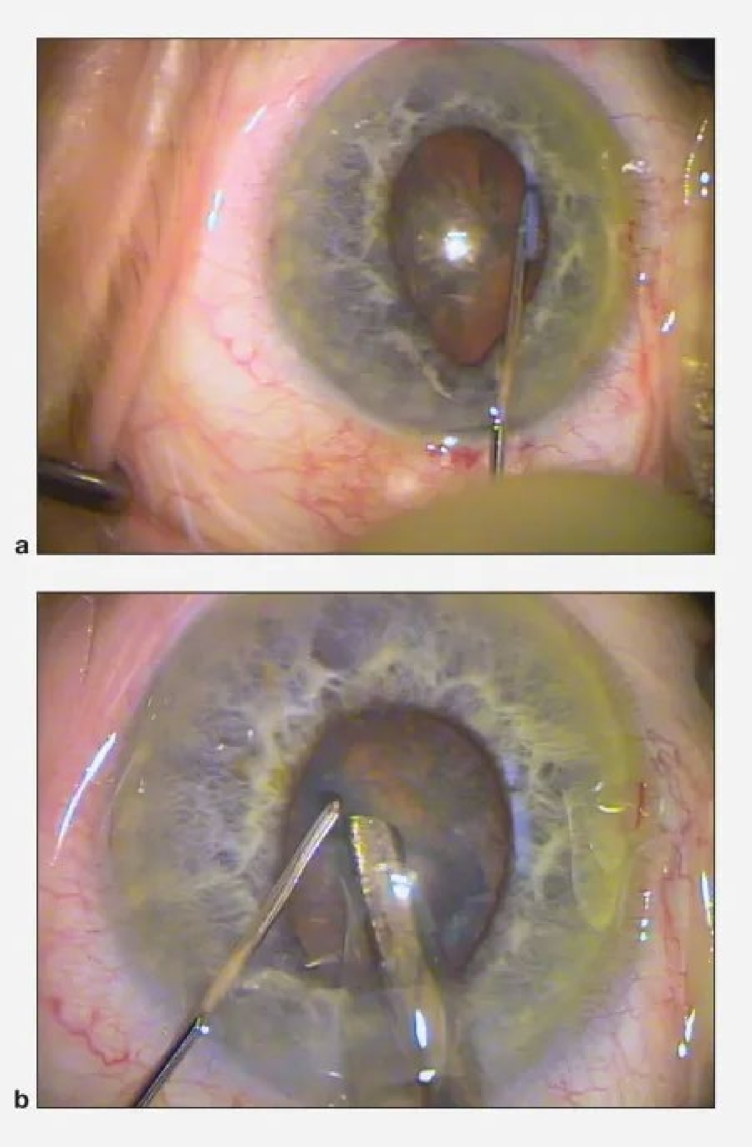

Along with tamsulosin, which medication is the most likely to cause this problem?

1, Lidocaine

2, Silodosin

3, Alfuzosin

4, Finasteride

Multimodal model (with images) answer: The images provided show a condition during what appears to be cataract surgery. The problem shown could be Intraoperative Floppy Iris Syndrome (IFIS), which is a complication that can occur during cataract surgery associated with the use of certain medications. Among the choices provided: Lidocaine is a local anesthetic and does not typically affect the iris during cataract surgery. Silodosin is a selective alpha-1 receptor antagonist used to treat benign prostatic hyperplasia (BPH) and is known to potentially cause IFIS. Alfuzosin is also an alpha-1 receptor antagonist used for BPH, which can cause IFIS. Finasteride is a medication that treats BPH by inhibiting the conversion of testosterone to dihydrotestosterone (DHT) and is not typically associated with IFIS. Considering tamsulosin, which is well known to be associated with IFIS, and given the choices, the most likely medication to also cause this problem would be:2, Silodosin

This is because both tamsulosin and silodosin are alpha-1 receptor antagonists and have a similar mechanism of action, which can lead to the occurrence of IFIS during cataract surgery.

**Case 3:** An example of misdiagnosis. A case of acute hydrops in keratoconus misdiagnosed as Acanthamoeba keratitis. The percentage for the general correct answer is 95.

https://www.aao.org/education/diagnose-this-player/diagnose-this-patient-with-contact-lens-complains-

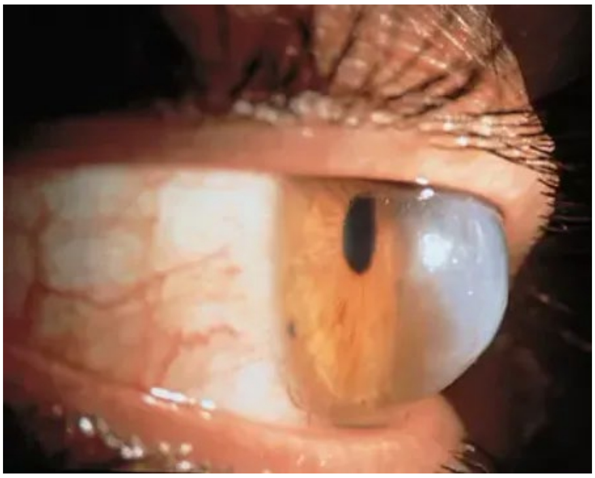

Question: A patient with keratoconus is fit with a rigid gas-permeable contact lens and is happy with the fit. Six months later, he calls your office and explains angrily that he saw his family practitioner for a red eye that he developed 3 days ago. The family doctor told the patient he had a serious corneal infection, prescribed antibiotics, and referred him to you for treatment. The lesion appears as shown. There is no epithelial defect, and the anterior chamber is deep and has only slight flare and a few cells. However, you note photophobia, tearing, conjunctival hyperemia, and vision decreased to bare finger-counting. What is the most likely diagnosis?

1, Corneal perforation

2, Bacterial keratitis

3, Acute corneal hydrops

4, Acanthamoeba keratitis

Multimodal model (with images) answer: The image shows a corneal lesion with no epithelial defect, and the patient is presenting with symptoms such as photophobia, tearing, conjunctival hyperemia, and significantly decreased vision. Considering these symptoms and the absence of an epithelial defect, along with the fact that the patient was wearing a rigid gas-permeable contact lens, which is a known risk factor, the most likely diagnosis from the given options would be: 4, Acanthamoeba keratitis.

Acanthamoeba keratitis is often associated with contact lens use, particularly if there has been exposure to contaminated water. This condition can present without an initial epithelial defect and can cause severe pain, photophobia, tearing, and red eye, which corresponds with the symptoms described. Bacterial keratitis typically would present with an epithelial defect and more purulent discharge. Acute corneal hydrops is associated with keratoconus but would present differently, and corneal perforation would have a significant epithelial defect and more severe anterior chamber reaction.

**Case 4:** An example successfully answered without giving a clear diagnosis.

A case of posterior polar cataract. The percentage for the general correct answer is 88.

https://www.aao.org/education/diagnose-this-player/diagnose-this-woman-with-lens-opacity

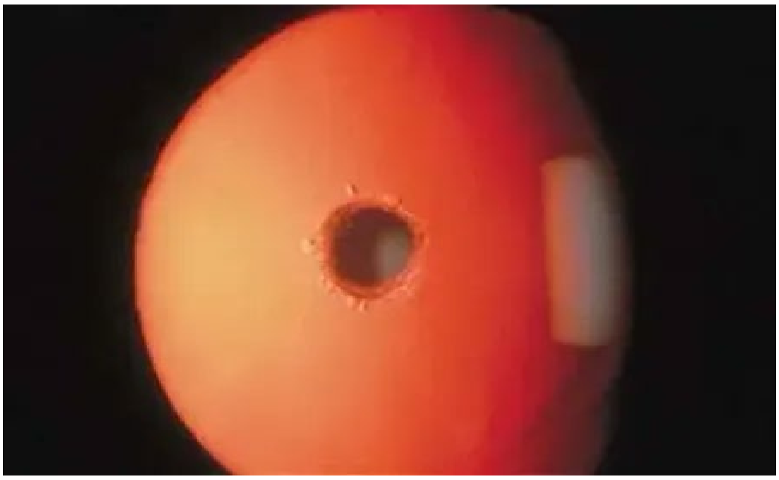

A woman in her 30s presents with 20/40 vision in both eyes correctable to 20/25 bilaterally and a lens opacity as shown. If the lens opacity becomes progressive, which one of the following complications is this patient's eye prone to develop?

1, Intraoperative capsular rupture

2, Intraoperative zonular dialysis

3, Postoperative choroidal detachment

4, Postoperative uveitis

Multimodal model (with images) answer: The image appears to show a lens with an opacity characteristic of a cataract. The most common complication that can occur due to progressive lens opacity, such as a cataract, particularly when it becomes dense and hard (known as a mature or hyper mature cataract), is intraoperative capsular rupture. This is because the lens capsule can become more brittle, and the cataract itself can be more challenging to remove. Therefore, the most likely complication that this patient's eye is prone to develop if the lens opacity becomes progressive is: Intraoperative capsular rupture.
